# What can be learned from viral co-detection studies in human populations

**DOI:** 10.1101/2023.06.17.23291541

**Authors:** Taylor Chin, Ellen F. Foxman, Timothy A. Watkins, Marc Lipsitch

## Abstract

When respiratory viruses co-circulate in a population, individuals may be infected with multiple pathogens and experience possible virus-virus interactions, where concurrent or recent prior infection with one virus affects the infection process of another virus. While experimental studies have provided convincing evidence for within-host mechanisms of virus-virus interactions, evaluating evidence for viral interference or potentiation using population-level data has proven more difficult. Recent studies have quantified the prevalence of co-detections using populations drawn from clinical settings. Here, we focus on selection bias issues associated with this study design. We provide a quantitative account of the conditions under which selection bias arises in these studies, review previous attempts to address this bias, and propose unbiased study designs with sample size estimates needed to ascertain viral interference. We show that selection bias is expected in cross-sectional co-detection prevalence studies conducted in clinical settings, except under a strict set of assumptions regarding the relative probabilities of having symptoms under different viral states. Population-wide studies that sample participants irrespective of their symptom status would meanwhile require large sample sizes to be sufficiently powered to detect viral interference, suggesting that a study’s timing, inclusion criteria, and the expected magnitude of interference are instrumental in determining feasibility.

## Overview of virus-virus interactions

Seasonal cocirculation of respiratory viruses may result in individuals being infected concurrently or sequentially with multiple pathogens. In this context, virus-virus interaction is often broadly defined as the phenomenon where concurrent or recent prior infection with one virus affects the infection process of another virus.^1^ Interactions may be classified as positive (synergistic) if infection with one virus promotes the presence of another virus or negative (antagonistic, competitive) if infection with one virus impedes the presence of another virus. An additional classification described in the literature distinguishes homologous and heterologous interference within negative virus-virus interactions based on whether two viruses are antigenically distinct.^1^

In a null model in which two viruses A and B transmit independently from one another, knowledge that an individual harbors one virus would provide no information about whether they harbor the other – that is,

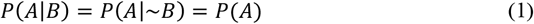

If *Equation (1)* is not true, there could be several explanations for an observed statistical association between viruses A and B under a causal inference framework^2^: (1) virus A promotes or inhibits virus B, (2) virus B promotes or inhibits virus A, (3) there is a common cause, *L*, of infection or noninfection with viruses A and B (also known in epidemiology as a confounder of the causal relationship between the infection processes of viruses A and B), or (4) the sample has been collected in a way that conditions on a common effect, *C*, of two variables – one of which is virus A or a variable associated with virus A, and the other is virus B or a variable associated with virus B (also known as selection bias or collider-stratification bias).

Whereas explanations (1) and (2) reflect interactions within an individual between the infection processes of two viruses, explanations (3) and (4) do not. Studies attempting to examine phenomena (1) or (2) should therefore ordinarily try to minimize bias from confounding or selection bias. The causal mechanisms underlying explanations (1) and (2) can be due to “biological/direct” mechanisms (*e*.*g*. antiviral role of activated interferon-stimulated genes)^3–9^, and/or “behavioral/indirect” mechanisms that lead to people infected with virus A not getting infected with virus B (*e*.*g*. they are more likely to stay at home when feeling unwell). Age and immunocompetence are examples of explanation (3), or common causes of the risk of infection of each of the two viruses, that have been discussed in the literature.^10,11^

With regard to explanation (4), various studies have attempted to ascertain virus-virus interactions at the population level using cross-sectional co-detection prevalence studies, aided by the development of multiplex PCR.^3,12–15^ In this study design, the expected proportion of co-detections of two viruses under the null model of independence is calculated as the product of the individual prevalence of the two viruses. That is, if the prevalence of virus A is *p*_*a*_ and prevalence of virus B is *p*_*b*_ in the population, then under the null hypothesis of no interaction, the expected proportion of the population infected with both viruses would be *p*_*a*_*p*_*b*_ in an unbiased sample of the population. If the observed number of co-detections is significantly less than this expected number, negative interaction, or viral interference, is inferred.

Key to this study design is the assumption that an unbiased sample of the population is used, that is, a sample in which the prevalence of each virus and of the two together is the same as that in the source population. Co-detection studies, however, are often conducted using clinical samples from people with acute respiratory illness (ARI) symptoms due to ease of surveillance in these populations. Selection bias, or collider-stratification bias, is a common methodological issue with this type of study.^2^ An intuitive way to think about the bias is that the sample of symptomatic patients presenting to the hospital with ARI symptoms is likely to have higher proportions of individuals with at least one of the respiratory viruses than the population from which it is drawn. It has therefore been correctly argued that finding a departure from independence in such a sample – a reduced proportion of co-infected compared to the expected proportion based on the prevalence of singly infected individuals – may reflect only this biased sampling and not necessarily any underlying antagonism in the population.^16–18^ *Table 1* summarizes different possible explanations for an observed statistical association between viruses A and B.

**Table 1.**
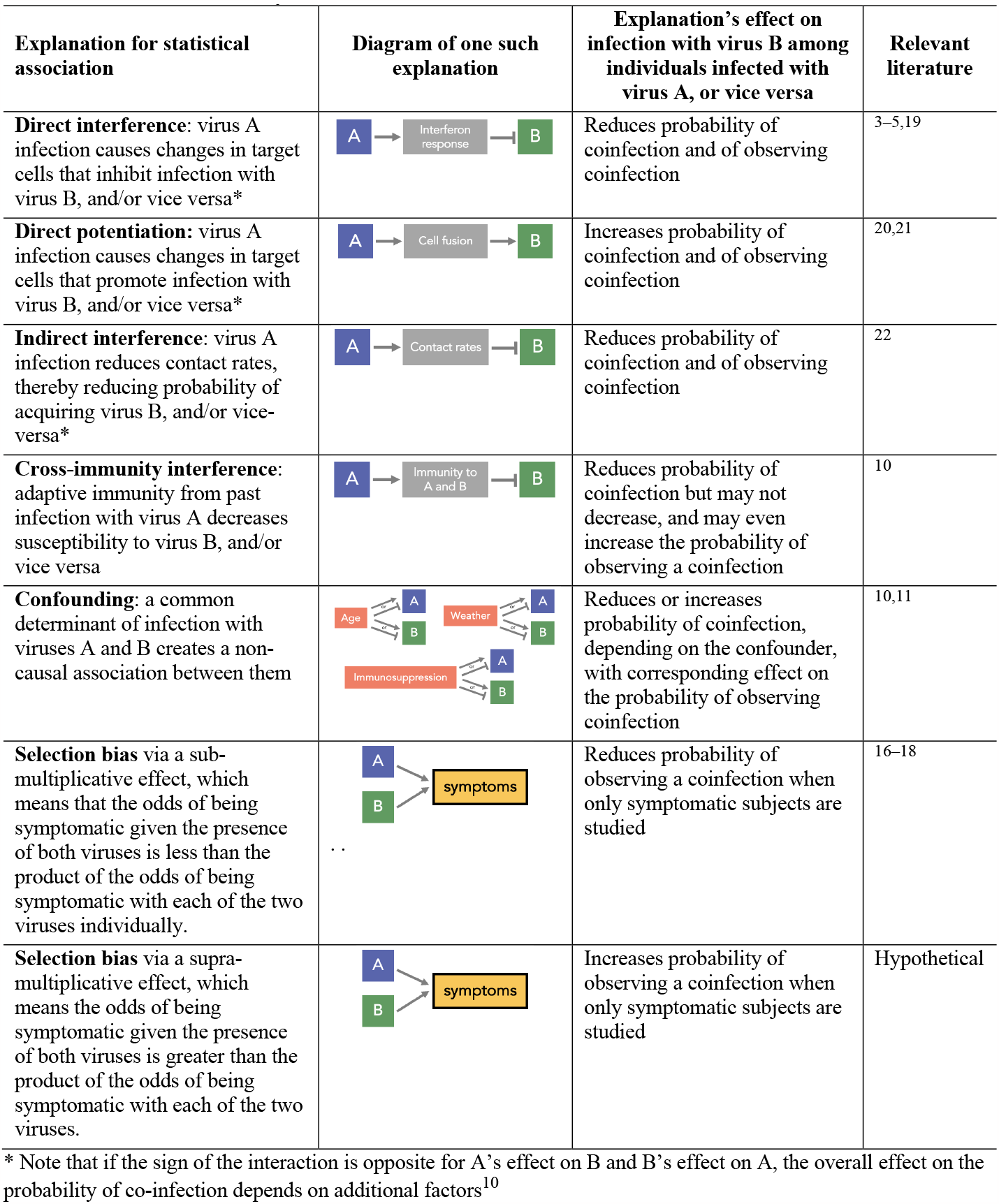
Possible explanations for an observed statistical association between the cross-sectional prevalence of two viruses (that is, *p*_*ab*_ ≠ *p*_*a*_*p*_*b*_, where *p*_*ab*_ is the prevalence of infection with both viruses simultaneously).

| Explanation for statistical association | Diagram of one such explanation | Explanation's effect on infection with virus B among individuals infected with virus A, or vice versa | Relevant literature |
| --- | --- | --- | --- |
| **Direct interference:** virus A infection causes changes in target cells that inhibit infection with virus B, and/or vice versa* |  | Reduces probability of coinfection and of observing coinfection | 3–5,19 |
| **Direct potentiation:** virus A infection causes changes in target cells that promote infection with virus B, and/or vice versa* |  | Increases probability of coinfection and of observing coinfection | 20,21 |
| **Indirect interference:** virus A infection reduces contact rates, thereby reducing probability of acquiring virus B, and/or vice versa* |  | Reduces probability of coinfection and of observing coinfection | 22 |
| **Cross-immunity interference:** adaptive immunity from past infection with virus A decreases susceptibility to virus B, and/or vice versa |  | Reduces probability of coinfection but may not decrease, and may even increase the probability of observing a coinfection | 10 |
| **Confounding:** a common determinant of infection with viruses A and B creates a non-causal association between them |  | Reduces or increases probability of coinfection, depending on the confounder, with corresponding effect on the probability of observing coinfection | 10,11 |
| **Selection bias** via a sub-multiplicative effect, which means that the odds of being symptomatic given the presence of both viruses is less than the product of the odds of being symptomatic with each of the two viruses individually. |  | Reduces probability of observing a coinfection when only symptomatic subjects are studied | 16–18 |
| **Selection bias** via a supra-multiplicative effect, which means the odds of being symptomatic given the presence of both viruses is greater than the product of the odds of being symptomatic with each of the two viruses. |  | Increases probability of observing a coinfection when only symptomatic subjects are studied | Hypothetical |

\* Note that if the sign of the interaction is opposite for A's effect on B and B's effect on A, the overall effect on the probability of co-infection depends on additional factors<sup>10</sup>

**Table 1.**
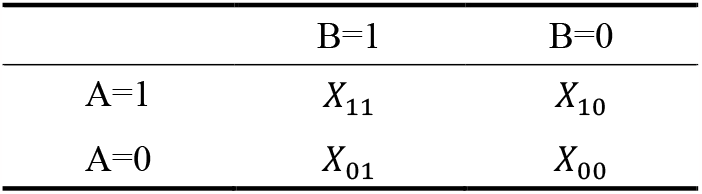
Population-level frequencies of having virus A and virus B. The directionality and strength of the OR would serve as indicators of virus-virus interaction, under a set of assumptions.

The SARS-CoV-2 pandemic has regenerated interest in studying virus-virus interactions because of their implications for disease forecasting and evaluation of interventions. This article will focus specifically on the selection bias issues associated with co-detection prevalence studies drawn from clinical setting populations given the study design’s simplicity and therefore widespread use in the absence of population-level cohort studies. Here, we first provide a quantitative account of the conditions under which selection bias arises in co-detection studies and its mechanistic interpretation, then discuss previous attempts to address this bias, and last, propose unbiased study designs with estimates of the sample sizes needed to ascertain viral interference. We focus on virus-virus interactions in the form of interference due to recent studies that have motivated this article, but the same arguments equally apply to the case of potentiation.

## Defining the selection bias issue in co-detection studies

Let *A* and *B* be Bernoulli random variables representing the presence or absence of viruses A and B, respectively. Then, let *X*_*AB*_indicate the number of individuals with viruses A and/or B in the population.

The population odds ratio (OR) estimated from *Table 2, OR*_*X*_, is defined as the ratio of the odds of having virus A in the presence of virus B, relative to the odds of having virus A in the absence of virus B, in the population:

**Table 2.**
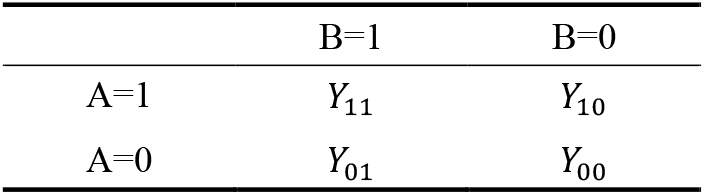
Sample-level frequencies of having virus A and virus B. The directionality and strength of the OR would serve as indicators of virus-virus interaction, if the sample is an unbiased sample of the population and a set of assumptions are met.

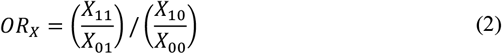

If *OR*_*X*_ < 1, the odds of having virus A in the presence of virus B is less than the odds of having virus A in the absence of virus B. This is symmetric, in that *OR*_*X*_ < 1 also indicates that the odds of having virus B in the presence of virus A is less than the odds of having virus B in the absence of A. Using the probability notation from the previous section, an alternative way to express a negative statistical association between viruses A and B is the prevalence ratio, 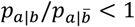, where the prevalence of virus A among individuals with virus B is 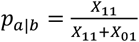 and the prevalence of virus A among individuals without virus B is 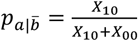.

In reality, *OR*_*X*_ is rarely if ever observed because only a sample, not the whole population, is assayed for viral infection in any particular study. In practice, that sample often consists of patients who report ARI symptoms and present to a clinical setting, where they receive molecular testing. Let *Y*_*AB*_ indicate the number of individuals with viruses A and/or B among patients reporting ARI symptoms in a clinical setting.

Similarly, the OR estimated from *Table 3, OR*_*Y*_, is defined as the ratio of the odds of having virus A in the presence of virus B, relative to the odds of having virus A in the absence of virus B, in the clinical sample:

**Table 3.**
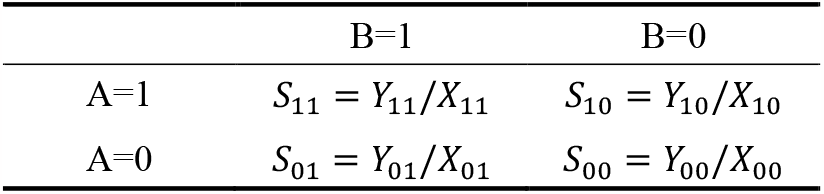
The probability that an individual with a given infection state is included in the sample of individuals with ARI symptoms.

|  | B=1 | B=0 |
| --- | --- | --- |
| A=1 | $S_{11} = Y_{11}/X_{11}$ | $S_{10} = Y_{10}/X_{10}$ |
| A=0 | $S_{01} = Y_{01}/X_{01}$ | $S_{00} = Y_{00}/X_{00}$ |

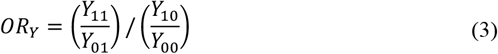

The values in *Table 3* are related to those in *Table 2* by the ratio of individuals with each virus state among ARI patients to that among the general population, *S*_*AB*_, which are summarized in *Table 4*. Conceptually, these probabilities represent the relative selection probabilities of people being included in the clinical sample based on their viral status.

**Table 4.**
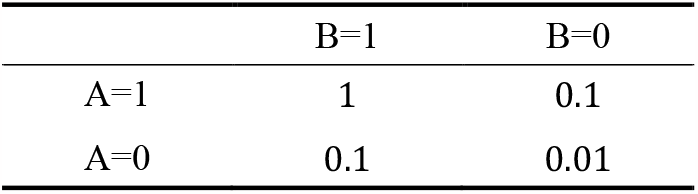
Example selection probabilities of having ARI symptoms given viral states A and B, where OR_S_ = 1 and there is therefore no selection bias

|  | B=1 | B=0 |
| --- | --- | --- |
| A=1 | 1 | 0.1 |
| A=0 | 0.1 | 0.01 |

The OR of this selection probability matrix, *OR*_*s*_, can be calculated by dividing the OR from the sample, *OR*_*Y*_, by the OR from the population, *OR*_*X*_.

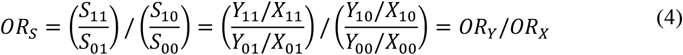

Greenland^23^ referred to *OR*_*S*_ as a selection-bias factor and pointed out that a corrected estimate, here the unbiased population-level OR, *OR*_*X*_, which is the estimand of interest, could be obtained by dividing the biased sample OR, *OR*_*Y*_, by the selection-bias factor, or the odds ratios of the selection probabilities:

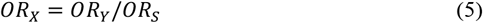

Knowledge of *OR*_*S*_ would be sufficient to obtain an unbiased estimate of *OR*_*X*_ – the population-level odds ratio, with all other assumptions holding for valid inference previously described. The key obstacle to this approach, however, is determining the selection probabilities in *Table 4* in the first place, as they are generally unknown.

Looking at *Equation (5)*, we can understand somewhat intuitively the circumstances under which selection bias can arise when using a symptomatic sample. As an example of a scenario involving two viruses that each cause a ten-fold increase in the probability that an individual has ARI symptoms, possible values for the selection probabilities of *Table 4* could be the following:

There would be no selection bias in this sample of symptomatic patients since *OR*, = 1; alternatively, having virus A raises one’s probability of having symptoms by the same multiplicative factor regardless of having virus B, and equivalently, having virus B raises one’s odds of having symptoms by the same multiplicative factor regardless of having virus A.

If the viruses under investigation are individually more likely to cause symptoms and the probability of having symptoms with one virus differs depending on whether one has the other virus, one possibility is that there is a sub-multiplicative effect of viruses A and B together causing symptoms relative to their individual effects:

In this scenario, if the population *OR*_*X*_ = 1, the sample-calculated OR is biased downward (*OR*_*Y*_ = *OR*_*S*_ *OR*_*X*_ = 0.17) compared to the population OR.

As a final example, having either virus A or virus B alone could not raise the probability of having symptoms by much relative to not having either, while having both viruses could greatly increase the probability of having symptoms:

In this case, the viruses have a synergistic effect on pathogenesis, so that their joint presence has a supra-multiplicative effect on the likelihood of having symptoms relative to the effect of each individual virus. The OR is biased upward (*OR*_*Y*_ = 1.67) in this scenario under the null.

As these examples illustrate, the OR estimated from the clinical sample, *OR*_*Y*_, is unaffected by selection bias only when *OR*_*S*_ = 1, and without knowledge of the selection probabilities, the magnitude of selection bias cannot be estimated. Previous commentaries have specifically pointed to the probability of having ARI symptoms among individuals without either virus, *S*_00_, as likely significantly lower than that of the other selection probabilities, which would induce selection bias.^16,17^ Since *S*_00_ is in the numerator of *OR*_*S*_ all else equal, *OR*_*S*_ will be smaller (and thus more likely to be less than 1) if *S*_00_ is small. The foregoing shows more precisely the conditions under which *OR*_*S*_ < 1.

The examples from *Tables 5-7* also demonstrate that a related research question of trying to infer synergy in pathogenesis based on individuals’ viral states using data from symptomatic individuals is affected by the same selection bias issue. In particular, observing *OR*_*Y*_ > 1 in a symptomatic sample could imply that the two viruses increase the probability of symptoms more than multiplicatively (*OR*_*S*_ > 1) but could also imply that they tend to occur together more often than expected under independence in the population (*OR*_*X*_ >1), for example if they tend to be common in the same demographic groups.

**Table 5.**
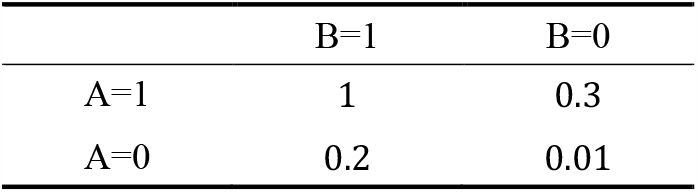
Example selection probabilities of having ARI symptoms given viral states A and B, where the OR_S_ < 1 (i.e. bias downward under the null) because of the sub-multiplicative effect of the viruses together causing symptoms

**Table 6.**
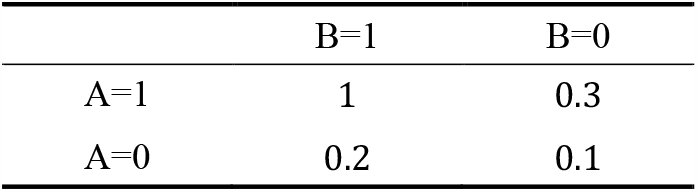
Example selection probabilities of having ARI symptoms given viral states A and B, where the OR_S_ > 1 (i.e. bias upward under the null) because of the supra-multiplicative effect of the viruses together causing symptoms

**Table 7.**
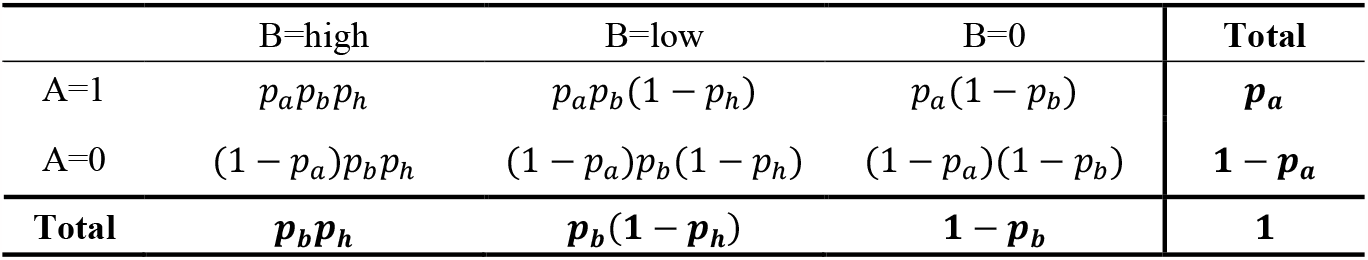
Population proportion frequencies of having virus A and virus B, where individuals with virus B are separated into high vs. low viral load groups

|  | B=high | B=low | B=0 | Total |
| --- | --- | --- | --- | --- |
| A=1 | $p_a p_b p_h$ | $p_a p_b (1 - p_h)$ | $p_a (1 - p_b)$ | $p_a$ |
| A=0 | $(1 - p_a) p_b p_h$ | $(1 - p_a) p_b (1 - p_h)$ | $(1 - p_a) (1 - p_b)$ | $1 - p_a$ |
| Total | $p_b p_h$ | $p_b (1 - p_h)$ | $1 - p_b$ | **1** |

### Related study designs and research questions

Experimental models for viral interference have shown that coinfection or sequential infection often results in decreased replication of virus B in the presence of virus A rather than a complete blockade of virus B.^3,4^ Examining viral loads in coinfections has therefore been proposed as a method to evaluate virus-virus interactions. Recently, using a sample of symptomatic individuals, Burstein *et al*. (2022) restricted consideration to individuals with virus B and compared the *abundance* of virus B among individuals with and without virus A, instead of comparing the presence and absence of virus B among individuals with and without virus A. This comparison involved estimating the average qPCR cycle threshold (Ct) value difference between monoinfected versus coinfected samples, adjusted for confounders.^17^ We show below that the same form of selection bias is likely to appear in this design. While Burstein *et al*. considered abundance of virus B as a continuous quantity, we illustrate for simplicity by dichotomizing virus B infected individuals as having high or low viral loads.

Consider expected proportions under the null hypothesis of no interaction in the population. Let *p*_*a*_ be the prevalence of virus A and *p*_*b*_ be the prevalence of virus B. Individuals with virus *B* are further separated into “high” and “low” viral load groups based on the probability of having high viral load given having virus *B, p*_*h*_. Here, a high viral load corresponds to low Ct values based on a cutoff value. The population proportion frequencies are summarized as the following:

The population odds ratio for any two columns of *Table 8* (or for the first two columns collapsed vs. the third, mimicking the presence/absence study design) is 1 under the null hypothesis. However, in a symptomatic population, the selection probabilities again enter the calculation.

**Table 8.** Selection probabilities, representing the probability of being in the sampled population having ARI symptoms, for each category of presence/absence of virus A and abundance of virus B.

|  | B=high | B=low | B=0 |
| --- | --- | --- | --- |
| A=1 | $S_{1,high}$ | $S_{1,low}$ | $S_{1,0}$ |
| A=0 | $S_{0,high}$ | $S_{0,low}$ | $S_{0,0}$ |

Since the analysis is restricted to individuals positive for virus B, to estimate the sample OR, element-wise multiplication is applied to the matrix values in *Tables* 8 and *9*, and the OR of the resultant matrix, excluding the *B* = 0 column, is calculated. The sample odds ratio would be 1 under the null only if:

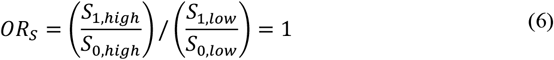

This modified study design therefore also ultimately relies on similar assumptions for the relative pathogenesis of viruses A and B and their joint probabilities of causing symptoms, but in this case, assumptions regarding the selection probabilities are made based on the presence/absence of virus A and the relative viral abundance of virus B. Although this example reduces the analysis by Burstein *et al*. (2022) into a binary example for their semi-quantitative viral load measure, if the selection bias issue is present in the binary case, it also applies to the case of a quantitative measure for viral load. A related limitation of this design is that it relies on a dose-response effect of the amount of B on A. If the interaction is so strongly inhibitory that a high viral load of B is incompatible with A, the design would miss this strongest of all possible interactions.

In summary, selection bias in co-detection prevalence studies arises because viral status and viral load likely affect the probability of symptoms and therefore the likelihood of individuals being included in the sample. The OR estimated from a clinical sample could be corrected to estimate the unbiased population-level OR if the selection probabilities (*i*.*e*. the probability of having ARI symptoms for different combinations of viruses A and B) were known, but they typically are not. This premise is important because selection bias is not an issue only in the case where the crossproduct of the selection probabilities equals one. Attempts to address this issue by using a semi-quantitative viral load measure for one virus are prone to the same selection bias issue. Likewise, use of symptomatic samples to assess whether viral infections are synergistic in causing symptoms falls prey to the same bias, in the other direction: an excess of dually infected individuals among symptomatic persons compared to single infection prevalence could be due to synergy in causing symptoms, or due to potentiation of infection by one virus when one is already infected with the other, or a combination of these two.

In the next section, we propose study designs and provide sample size estimates for studies not conditioned on symptom status that may be used to ascertain virus-virus interactions.

### Study designs to measure virus-virus interactions

Human challenge studies have been used for hundreds of years to garner greater scientific understanding of viral lifecycles and pathogenesis. In human challenge studies, volunteers are intentionally infected with a pathogen at a safe infectious dose and isolated in a quarantine unit during their infection, where they receive close and continuous medical monitoring.^24^ To our knowledge, human challenge studies have yet to be conducted to study virus-virus interactions. It is plausible that a study could be designed such that volunteers are randomized to be inoculated with either one virus or mock inoculation, and a few days, all volunteers are inoculated with a different challenge virus. Detailed information on symptoms progression, viral kinetics, and immune correlates of protection could be collected. This design would be analogous to the experimental studies that have investigated virus-virus interactions using organoid models^3–6,8,9^ and animal models,^7,25^ and it could help elucidate understanding of direct, within-host mechanisms of interaction, but not indirect mechanisms. Human challenge studies, however, are also associated with significant ethical considerations and require weighing prospective benefits to society relative to the higher risks posed to participants.

As another study design option, the effect of co-infection with respiratory viruses on a standardized attenuated challenge virus could be examined. A Phase 4 immunogenicity study of a trivalent live attenuated influenza vaccine (LAIV) in The Gambia recruited clinically well children and found that children with asymptomatic respiratory viruses had upregulated mucosal interferon responses, which correlated with reduced replication of live attenuated influenza viruses post-challenge, compared to children without viral infections at baseline.^26^ Relatedly, a study of children with cystic fibrosis and their siblings found decreased replication of a live attenuated influenza vaccine strain in participants who tested positive for another respiratory virus before vaccination.^27^

Another valid approach to assess virus interactions in humans would be observational studies that sample from populations in a way that does not condition on symptoms. This would involve population-based community studies, where nasopharyngeal swab samples are collected weekly from all participants regardless of symptoms. Moreover, to minimize confounding, ideally the participant age group and the time window of sample collection should be as narrow as possible, though these factors could also be adjusted for in the analysis, as we discuss below. To ensure specimen collection is done irrespective of symptoms, participants could collect specimen samples at home, and the samples could be retrieved every few days by study administrators for analysis. The OR estimated from the sample population in this case may serve as an estimate of the population unimpeded by selection bias. Self-reported symptom status at the time of participants’ sample collection could also be collected every week. This information on symptoms could then be used to estimate the selection probabilities of *Table 4*, and ORs estimated from clinical samples could be adjusted using *Equation (5)*. Longitudinal community surveillance studies using similar study designs have been conducted in New York City^28^, Utah^29,30^, and Australia^31^ and have revealed prevalence values for low pathogenicity viruses that are higher than previously thought. Also, certain populations have been sampled in this way during COVID-19 and samples from these studies may be among the largest sample sets ever collected in this way.

While large community-based sampling studies may represent a methodologically straight-forward approach to collecting unbiased data, they are expensive and have logistical challenges, such as the collection of specimen samples from volunteers. Conducting studies in settings where surveillance is already established and routinely performed, like in hospitals and clinical settings, therefore remains attractive, if individuals can be sampled irrespective of their symptom status. For example, collecting samples from individuals admitted to the ER for reasons unrelated to ARI symptoms could generate an unbiased study population in this setting, if one assumes that these individuals have the same prevalence of respiratory symptoms as the overall population.

To provide a sample size estimate for co-detection studies, we use possible virus-virus interaction between three pairs of viruses as examples. Based on estimates of children <10 years old from a community-based sampling study in New York City, the average prevalence over two winter seasons from October to February 2016-2017 and 2017-2018 was ∼18% for rhinovirus (RV), ∼4% for adenovirus, ∼10% for coronavirus, and ∼2% for influenza.^28^ In a hypothesis testing framework for two viruses, virus A and virus B, we would test the null hypothesis*H*_0_: *OR* = 1 and alternative hypothesis *H*_0_: *OR* ≠ 1, using the notation:

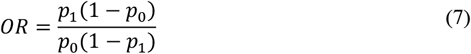

where *p*_0_ is the probability of an individual having virus B given they do not have virus A and *p*_%_ is the probability of an individual having virus B given they have virus A. The total sample size required, *n*, would then be calculated based on a two-sided hypothesis test for an OR estimated from a cross-sectional study^32^:

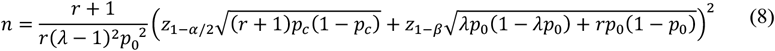

where

- *r* is the ratio of the number of individuals who have virus A to the number of individuals without virus A
- *λ* is the ratio of the prevalence of the outcome (virus B) in the exposed (virus A positive) to the prevalence of the outcome in the unexposed (virus A negative); that is, *p*_1_/*p*_0_
- *p*_*c*_ is the average of the prevalence of the outcome (virus B) in the exposed and unexposed; 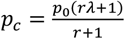
- *α* is the Type I error rate
- *β* is the Type II error rate

Using an example of RV as virus A and influenza virus as virus B, we assume *α* = 0.05, *β* = 0.2 for 80% power, *p*_0_ = 0.02, which is the prevalence of influenza virus in this population, and *r* = 0.175/0.825 ≈ 0.21, based on the prevalence of RV in the population. Using a range of OR values, which are used to calculate *p*_1_, the estimated required total sample size to detect viral interference between RV and influenza in children during a winter season would fall between approximately 1,760 participants for OR = 0.05 and 260,000 for OR = 0.9 (**Figure 1**). For RV and coronavirus, the range is between approximately 300 to 56,000 participants based on the expected OR. The examination of two low prevalence viruses like adenovirus and influenza would meanwhile require between approximately 7,000 and 1,000,000 participants depending on the OR.

**Figure 1.**
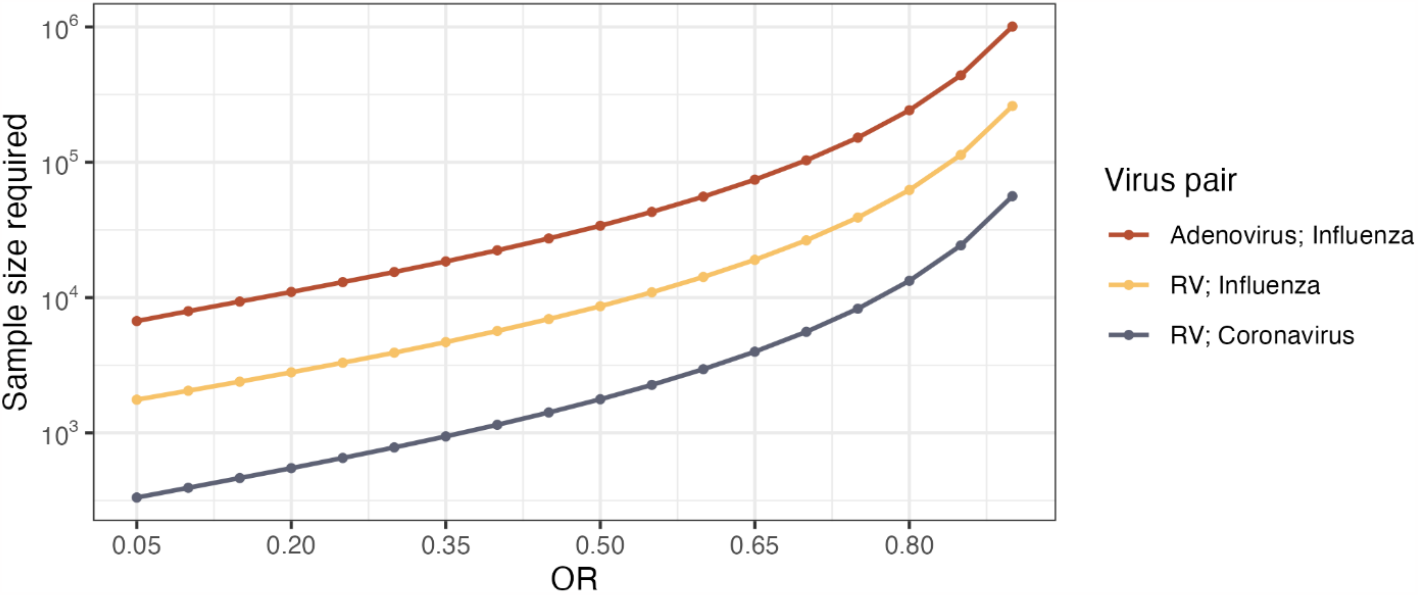
Estimated required total sample size (shown on log scale) needed to observe ORs between 0.05 and 0.9 for three example virus pairs in cross-sectional, co-detection prevalence studies.

These sample size estimates are lower bound estimates since the analysis would also need to adjust for confounding. Important confounders to consider include the age of participants and the time period over which samples are collected based on the study design and population. Additionally, if multiple study sites are used, the geographic region of the study site and specimen analysis method would also be potential confounders. These high sample size estimates indicate that studying viral interference using large population-based sampling studies may be most feasible if the magnitude of viral interference is believed to be significant and if the studies are conducted among highly susceptible populations like children during high viral circulation weeks to maximize the likelihood of capturing co-detections.

## Conclusion

The SARS-CoV-2 pandemic and its effect on the dynamics of other respiratory pathogens have spurred scientific interest in virus-virus interactions. Interest in virus-virus interactions also increased during the 2009 A(H1N1) pandemic, as ecological reports from several European countries suggested that high RV transmission during the summer delayed the onset of A(H1N1) outbreaks until late fall.^33,34^ Various studies have been conducted using cell culture and animal models to explore direct/biological mechanisms of viral interference.^3–9,19^ While these experimental studies have been instrumental in elucidating within-host mechanisms for viral interference, observing these mechanisms’ effects at the population level has been more elusive.

At the population level, studies have used time-series analyses and co-detection prevalence studies to ascertain virus-virus interactions that reflect a combination of direct and indirect, or behavior-driven, mechanisms.^12,30,31,35–37^ Modeling studies have also been conducted to show that using cross-sectional prevalence studies to infer the impact of multivalent vaccines on type replacement may lead to biased conclusions.^10,38^

Here, we focused on selection bias issues that arise when using clinical data from symptomatic patients in co-detection studies, which is a common practice due to the expense and logistic difficulties of conducting community-wide sampling studies. A key challenge with this study design is selecting individuals irrespective of their symptom status. We provided a mechanistic interpretation of the selection bias that arises in these studies and discussed the conditions under which it would be expected. Community-based viral sampling studies and clinical setting-based studies conducted among individuals being admitted and tested for reasons unrelated to ARI symptoms are two possibilities for unbiased study designs. The large sample sizes estimated to be required for these studies suggest that many current efforts are underpowered to detect virus-virus interactions. Given the implications of virus-virus interactions on disease forecasting and the design and implementation of public health interventions, there is a scientific need to ascertain possible interference and its effects using unbiased designs.

## Data Availability

All data produced in the present work are contained in the manuscript.

## References

1 Piret J, Boivin G. Viral Interference between Respiratory Viruses. Emerg Infect Dis 2022; 28: 273–81.

2 Hernán MA, Hernández-Díaz S, Robins JM. A structural approach to selection bias. Epidemiol Camb Mass 2004; 15: 615–25.

3 Wu A, Mihaylova VT, Landry ML, Foxman EF. Interference between rhinovirus and influenza A virus: a clinical data analysis and experimental infection study. Lancet Microbe 2020; 1: e254–62.

4 Cheemarla NR, Watkins TA, Mihaylova VT, et al. Dynamic innate immune response determines susceptibility to SARS-CoV-2 infection and early replication kinetics. J Exp Med 2021; 218: e20210583.

5 Geiser J, Boivin G, Huang S, et al. RSV and HMPV Infections in 3D Tissue Cultures: Mechanisms Involved in Virus-Host and Virus-Virus Interactions. Viruses 2021; 13: 139.

6 Essaidi-Laziosi M, Alvarez C, Puhach O, et al. Sequential infections with rhinovirus and influenza modulate the replicative capacity of SARS-CoV-2 in the upper respiratory tract. Emerg Microbes Infect 2022; 11: 412–23.

7 Oishi K, Horiuchi S, Minkoff JM, tenOever BR. The Host Response to Influenza A Virus Interferes with SARS-CoV-2 Replication during Coinfection. J Virol 2022; 96: e00765–22.

8 Van Leuven JT, Gonzalez AJ, Ijezie EC, et al. Rhinovirus Reduces the Severity of Subsequent Respiratory Viral Infections by Interferon-Dependent and -Independent Mechanisms. mSphere 2021; 6: e0047921.

9 Dee K, Schultz V, Haney J, Bissett LA, Magill C, Murcia PR. Influenza A and respiratory syncytial virus trigger a cellular response that blocks severe acute respiratory syndrome virus 2 infection in the respiratory tract. J Infect Dis 2022; : jiac494.

10 Man I, Wallinga J, Bogaards JA. Inferring Pathogen Type Interactions Using Cross-sectional Prevalence Data: Opportunities and Pitfalls for Predicting Type Replacement. Epidemiol Camb Mass 2018; 29: 666–74.

11 Hamelin FM, Allen LJS, Bokil VA, et al. Coinfections by noninteracting pathogens are not independent and require new tests of interaction. PLOS Biol 2019; 17: e3000551.

12 Price OH, Sullivan SG, Sutterby C, Druce J, Carville KS. Using routine testing data to understand circulation patterns of influenza A, respiratory syncytial virus and other respiratory viruses in Victoria, Australia. Epidemiol Infect 2019; 147: e221.

13 Mackay IM, Lambert SB, Faux CE, et al. Community-Wide, Contemporaneous Circulation of a Broad Spectrum of Human Rhinoviruses in Healthy Australian Preschool-Aged Children During a 12-Month Period. J Infect Dis 2013; 207: 1433–41.

14 Greer RM, McErlean P, Arden KE, et al. Do rhinoviruses reduce the probability of viral co-detection during acute respiratory tract infections? J Clin Virol 2009; 45: 10–5.

15 Achten NB, Wu P, Bont L, et al. Interference Between Respiratory Syncytial Virus and Human Rhinovirus Infection in Infancy. J Infect Dis 2017; 215: 1102–6.

16 Nordén R, Lindh M, Nilsson S, Westin J. Viral interference cannot be concluded from datasets containing only symptomatic patients. Lancet Microbe 2021; 2: e9.

17 Burstein R, Althouse BM, Adler A, et al. Interactions among 17 respiratory pathogens: a cross-sectional study using clinical and community surveillance data. 2022; : 2022.02.04.22270474.

18 Wu A, Mihaylova VT, Landry ML, Foxman EF. Viral interference cannot be concluded from datasets containing only symptomatic patients – Authors’ reply. Lancet Microbe 2021; 2: e10.

19 Chan KF, Carolan LA, Korenkov D, et al. Investigating Viral Interference Between Influenza A Virus and Human Respiratory Syncytial Virus in a Ferret Model of Infection. J Infect Dis 2018; 218: 406–17.

20 Goto H, Ihira H, Morishita K, et al. Enhanced growth of influenza A virus by coinfection with human parainfluenza virus type 2. Med Microbiol Immunol (Berl) 2016; 205: 209–18.

21 Bai L, Zhao Y, Dong J, et al. Coinfection with influenza A virus enhances SARS-CoV-2 infectivity. Cell Res 2021; 31: 395–403.

22 Rohani P, Earn DJ, Finkenstädt B, Grenfell BT. Population dynamic interference among childhood diseases. Proc R Soc B Biol Sci 1998; 265: 2033–41.

23 Greenland S. Basic methods for sensitivity analysis of biases. Int J Epidemiol 1996; 25: 1107– 16.

24 Human challenge trials for vaccine development: regulatory considerations, Annex 10, TRS No 1004. 2017; published online Jan. https://www.who.int/publications/m/item/human-challenge-trials-for-vaccine-a10-trs-no-1004 (accessed Jan 7, 2023).

25 Laurie KL, Guarnaccia TA, Carolan LA, et al. Interval Between Infections and Viral Hierarchy Are Determinants of Viral Interference Following Influenza Virus Infection in a Ferret Model. J Infect Dis 2015; 212: 1701–10.

26 Costa-Martins AG, Mane K, Lindsey BB, et al. Prior upregulation of interferon pathways in the nasopharynx impacts viral shedding following live attenuated influenza vaccine challenge in children. Cell Rep Med 2021; 2: 100465.

27 Boikos C, Papenburg J, Martineau C, et al. Viral interference and the live-attenuated intranasal influenza vaccine: Results from a pediatric cohort with cystic fibrosis. Hum Vaccines Immunother 2017; 13: 1254–60.

28 Galanti M, Birger R, Ud-Dean M, et al. Longitudinal active sampling for respiratory viral infections across age groups. Influenza Other Respir Viruses 2019; 13: 226–32.

29 Byington CL, Ampofo K, Stockmann C, et al. Community Surveillance of Respiratory Viruses Among Families in the Utah Better Identification of Germs-Longitudinal Viral Epidemiology (BIG-LoVE) Study. Clin Infect Dis 2015; 61: 1217–24.

30 Adler FR, Stockmann C, Ampofo K, Pavia AT, Byington CL. Transmission of rhinovirus in the Utah BIG-LoVE families: Consequences of age and household structure. PLoS ONE 2018; 13: e0199388.

31 Takashima MD, Grimwood K, Sly PD, Lambert SB, Ware RS. Interference between rhinovirus and other RNA respiratory viruses in the first 2-years of life: A longitudinal community-based birth cohort study. J Clin Virol 2022; 155: 105249.

32 Woodward M. Epidemiology: Study Design and Data Analysis, Third Edition, 3rd edn. New York: Chapman and Hall/CRC, 2014 DOI:10.1201/b16343.

33 Casalegno JS, Ottmann M, Duchamp MB, et al. Rhinoviruses delayed the circulation of the pandemic influenza A (H1N1) 2009 virus in France. Clin Microbiol Infect 2010; 16: 326–9.

34 Linde A, Rotzén-Östlund M, Zweygberg-Wirgart B, Rubinova S, Brytting M. Does viral interference affect spread of influenza? Eurosurveillance 2009; 14: 19354.

35 Nickbakhsh S, Mair C, Matthews L, et al. Virus–virus interactions impact the population dynamics of influenza and the common cold. Proc Natl Acad Sci 2019; 116: 27142–50.

36 Shrestha S, Foxman B, Weinberger DM, Steiner C, Viboud C, Rohani P. Identifying the Interaction Between Influenza and Pneumococcal Pneumonia Using Incidence Data. Sci Transl Med 2013; 5: 191ra84–191ra84.

37 Shrestha S, King AA, Rohani P. Statistical Inference for Multi-Pathogen Systems. PLOS Comput Biol 2011; 7: e1002135.

38 Durham DP, Poolman EM, Ibuka Y, Townsend JP, Galvani AP. Reevaluation of Epidemiological Data Demonstrates That It Is Consistent With Cross-Immunity Among Human Papillomavirus Types. J Infect Dis 2012; 206: 1291–8.

